# Artificial intelligence tool for the study of COVID-19 microdroplet spread across the human diameter and airborne space

**DOI:** 10.1101/2022.06.01.22275872

**Authors:** Hesham H. Alsaadi, Monther Aldwairi, Faten Yasin, Sandra C. P. Cachinho, Abdullah Hussein

## Abstract

The 2019 novel coronavirus (SARS-COV2 / COVID-19), with a point of origin in Wuhan, China, has spread rapidly all over the world. It turned into a raging pandemic wrecking havoc on health care facilities, world economy and affecting everyone’s life to date. With every new variant, rate of transmission, spread of infections and the number of cases continues to rise at an international level and scale. There are limited reliable researches that study microdroplets spread and transmissions from human sneeze or cough in the airborne space. In this paper, we propose an intelligent technique to visualize, detect, measure the distance of the spread in a real-world settings of microdroplet transmissions in airborne space, called ”COVNET45”. In this paper, we investigate the microdroplet transmission and validate the measurements accuracy compared to published researches, by examining several microscopic and visual images taken to investigate the novel coronavirus (SARS-COV2 / COVID-19). The ultimate contribution is to calculate the spread of the microdroplets measurements precisely with graphical presentation.

## Introduction

The 2019 novel coronavirus (SARS-COV2 / COVID-19) is an infectious respiratory disease that shares the same means and mechanisms of transmission as influenza [1]. Infectious disease transmission through respiratory secretion can spread via:

- Droplets – large particles (¿5 µm) that travel under 1m.
- Aerosols – smaller particles (¡5 µm) that travel over 1m.
- Contact - with objects that have been infected with droplets and then touching the eyes, nose, or mouth [2].

The airborne transmission does not require direct contact between infected and susceptible individuals; that virus causing COVID-19 might spread via droplets, e.g. when a person coughs. One can inhale microscopic aerosol particles consisting of the reliable residual components of evaporated respiratory droplets, which are tiny enough to remain airborne for hours. The aerosolized SARS-CoV-2 can remain viable and infectious in aerosols for hours and on certain surfaces up to days. But how do asymptomatic infected individuals generate aerosols? The normal acts of breathing or speaking could emit large quantities of aerosol particles. They are of approximately 1 µm to 500 µm in diameter but big enough to carry the viruses. [3].

Furthermore, some individuals are ”super speech emitters,” having the ability to emit more aerosol particles than others. For example, a 10 minutes conversation with an infected, asymptomatic super emitter talking in an average volume, could generate an invisible ”cloud” of approximately 6,000 aerosol particles that could potentially be inhaled by the other party [4] [5] [6]. It is then vital to understand the dynamics of the spread of cough and breathing particles of different sizes.

Gralton summarized the size of coughed particles from a large number of studies and concluded that the size of cough-generated particles ranged from 0.1–100 µm. [7]. To study the stability of COVID-19, some research groups work involved artificially generated, and aged aerosols using a nebulizer and maintaining it suspended in the air with a Goldberg drum. The viruses in different environmental conditions would remain viable in aerosols throughout the experiment, 3 hours; however, the virus titer significantly reduced after 3 hours. [8].

At present, there are available vaccines and are now produced and administered to individuals to prevent serious side effects of the virus they do not stop the spread [12]. Considering that its still a highly contagious virus, mask mandates in closed areas remain the main measure to prevent any droplet inhalation. Therefore, the best way of prevention is avoiding exposure, and the implementation of personal protective measures, which remain vital to mitigate and control the pandemic. The Centres for Disease Control and Prevention (CDC) & The World Health Organization (WHO) recommend that people avoid touching their faces [13]. The recommendation is to wear eye and face masks to minimize the risk of transmission. [9] [10].

However, wearing masks may create a false sense of security, and the subject should not neglect all the other essential preventative measures: hand hygiene, social distance, respiratory etiquette, and self-isolation if in close contact or exhibiting any symptoms [10]. Different masks types have different protection effectiveness and different breath-abilities. N95, or surgical masks, have higher protection efficiency of more than 90% against particle of 0.3-4.5 µm and reach about 100% for particles pf larger than 4.5 µm. It was shown that wearing a surgical face mask could prevent or reduce transmission of human coronaviruses and influenza viruses from symptomatic individuals. However, as the pandemic continues to claim lives, and calls for relaxing mask mandates such as lifting the restrictions in outdoor and open spaces, the need for masks indoors remains a must [11].

In this paper, we propose an intelligent technique to visualize, detect, measure the distance of the spread in a real-world settings of microdroplet transmissions in airborne space. The optimal goal of this paper is to: a) Develop a novel approach to optimize images with measuring the distance of microdroplets spread in a real-world scale of microdroplet transmissions in airborne space; we name it ”COVNET45” b) Provide an optimization technique via the utilization of state-of-the-art tools.

The main contributions of this work are:

- The proposed approach is to generate high accuracy measurement results by optimizing the collected images and their characterization.
- The ”COVNET45” tool combines strong capabilities of optimizing the detection as well as accurately measuring spread with the use of different scale system nominal, ordinal, interval and ratio.
- The proposed approach is generic in a sense, that is not limited to present only a small scale of microdroplets transmissions but also to measure the diameter of spread in any space using 2d graphical images with capabilities of analyzing, optimizing and detecting various semantic marks.

The rest of the paper is organized as follows. Section 2 discusses the existing related work and proposed methods for measurements systems in the literature. In Section 3, we describe the methodology, proposed approach and illustrate the workings of the COVNET45 tool. In Section 4, we show the implementation details and experimental results as well as discuss COVNET45 limitations. Section 5 concludes the paper.

### RELATED WORK

COVID-19 is a highly transmissible infectious respiratory illness that shares the routes and means of transmission with influenza. The largest size of microdroplets ¿5 µm can travel under 1m, while the smaller microdroplet particles ¡5 µm in the area can go over 1m in the distance.

Several studies have indicated that identifying the leading causes of the spread of COVID-19 may be through the transferal of infinitely small particles through the air. Proposed research used a background oriented schlieren technique to investigate the airflow ejected by a person quietly and heavily breathing, and coughing. They tested the effectiveness of different face covers including FFP2 and FFP1 masks, a respirator, a surgical and a hand-made mask, and two types of face shields, which simulate an aerosol-generating procedure demonstrating the extent of aerosol dispersion [24]. The study concludes that all face covers except the respirator, allow a reduction of the front flow through jet by more than 90%. Although the results sound promising the experimental setup does not reveal the absolute maximum distance that a virus-laden fluid particle can travel, nor how the concentration of these particles varies spatially and temporally [19].

Another proposed study described a solution for rapid detection of COVID-19, using genotypic testing for SARS-Cov-2 virus in nasopharyngeal. The study demonstrated fluorescent microscopy, and CT scans images of COVID-19 patient along with whole patient blood and platelet-poor plasma. The study concluded that micro clots can be detected in the native plasma of COVID-19 patient, and in particular that such clots are amyloid in nature as judged by a standard fluorogenic stain. Moreover, the study found that the plasma of COVID-19 patients carries a massive load of preformed amyloid clots. These clots imaged with TEG to provide a rapid, early detection test for clotting severity in such patients. The limitation of the study is the indication of the virus spread within the micrographic images of the plasma test [20].

Another study investigated the speech droplets generated by asymptomatic carriers of severe acute respiratory syndrome coronavirus 2 (SARS-CoV-2). The research considered that it is likely the mode of disease transmission. Highly sensitive laser light scattering observations have revealed that loud speech can emit thousands of oral fluid droplets per second. In a closed, stagnant air environment, they disappear from the window of view with time constants in the range of 8 to 14 min, which corresponds to droplet nuclei of ca. 4 µm diameter, or 12-to 21-µm droplets before dehydration. These observations confirm that there is a substantial probability that normal speaking causes airborne virus transmission in confined environments. The study cannot provide validation accuracy of the size of the diameter of the droplet using the light scattering observation of airborne speech droplet nuclei [3].

A recent study demonstrated the potential of microdroplet infection transmission in movement, such as walking fast, running and cycling can increase the infection spread to other individuals in public areas. The aerodynamics study investigated whether a first-person moving nearby a second person at 1.5 m distance or beyond could cause droplet transfer. The study demonstrated a CFD simulation of droplet dispersion around two walkers with exhaling velocity of 2.5 m/s relative to the movement of the walker/runner, representing moderately deep breathing. Moreover, the research experiments illustrated water droplets presented as (saliva) released at a total flow rate of 1×10-14 mg/s with a Rosin-Rammler droplet distribution with a minimum diameter of 40 µm, an average diameter of 80 µm and a maximum diameter of 200 µm. The study concluded that for fast walking at 4 km/h the spread distance is about 5 m and for running at 14.4 km/h, the spread distance is about 10 m [18].

Similar research explained the role of aerosols in the transmission of COVID-19, the research described two possible mode of COVID-19 aerosols transmission; a) during a sneeze or a cough, ”droplet sprays” of virus-laden respiratory tract fluid, typically greater than 5 µm in diameter, impact directly on a susceptible individual and b) alternatively, a susceptible person can inhale microscopic aerosol particles consisting of the residual solid components of evaporated respiratory droplets, which are tiny enough (¡5 µm) to remain airborne for hours [14].

Some studies used practical simulation to study the method of transmission by using robots to study the molecular splash of droplets spread to demonstrate the diffusion process indicating spread up to 8 feet of molecules through the air for the uncovered individual without the mask [17]. Moreover, the study used artificial materials of droplets simulated as real microdroplets from a human. Very concise recent studies have illustrated that the COVID-19 virus distribution process is one of its principal causes of widespread transmission through the air, as the amount of spread caused by an individual varies by the size of the droplets executed by the human body, which differs from sneezing or coughing.

The research studies, mentioned earlier, adopted different techniques for scaling and measuring microdroplets infection transmissions either by simulating graphical representation of microscopic images or by visualizing the spread using CFD examination. Due to the complexity of authenticating results in each research investigating the virus spreads through patient cough or sneeze, releasing respiratory droplets that carry the virus, many of the discussed studies lack in providing evidentiary tools to validate the results explicitly obtained through measurements and scaling droplets. Some studies also supported only specific data formats (CT scans, micrographic/microscopic images, CFD images, video graphics), and even when handling multiple formats, they cannot compare or validate results because of the use and restrictions provided by the investigation technology used. The fact that microdroplets patterns are ideally identifiable and recognizable makes it challenging to measure with precise size even when using automated scaling systems implemented within the technology. Finally, extensive types of scales formats in measuring accurate microdroplets or any microscopic details within the hierarchy is challenging, leaves the door open for more optimized and investigative tools to explore more and authenticate studies about the infectious disease.

Next, we explain the proposed methodology and structure of the proposed tool. COVNET45 optimization and measurements using graphical GUI representation that generates unique insights and accurate results.

## METHODOLOGY

COVNET45 uses images as a data source for processing and optimizing. In this section, first, we show the level relational paradigm of microscopy images. Second, we explain the process of optimizing microscopy images using multiple machine learning algorithms. Third, we use a scale calculation method to validate the results. Finally, we export and visualize the optimized solution.

### Objects,Events and Change Managements

In this section, we detail the process of managing microscopic images of COVID-19 virus using electron microscope device, how the COVID-19 virus distributed and organized in each cell and broken into objects in a single microscopic image file. In COVNET45, we consider that single spherical virus particles contain black dots of COVID-19 virus as an *object* in a single microscopic image, and the possible movements/diameter in size of a single spherical virus particle as *events*. Fig **??** shows a basic framework using a sample image thin-section electron microscopic image of SARS-CoV-2, the causative agent of COVID-19. Spherical virus particles contain black dots, which are cross-sections through the viral nucleocapsid. In the cytoplasm of the infected cell, clusters of particles are found within the membrane-bound cisternae of the rough endoplasmic reticulum/Golgi area.

Figure 1.B.(I) shows a sample image of thin-section electron microscopic image of SARS-CoV-2, the causative agent of COVID-19. Spherical virus particles contain black dots, which are cross-sections through the viral nucleocapsid. In the cytoplasm of the infected cell, clusters of particles are found within the membrane-bound cisternae of the rough endoplasmic reticulum/Golgi area. In COVNET45, the red markers represent the two objects X and Y, are shown by figure 1.B.(II).

**Fig 1.**
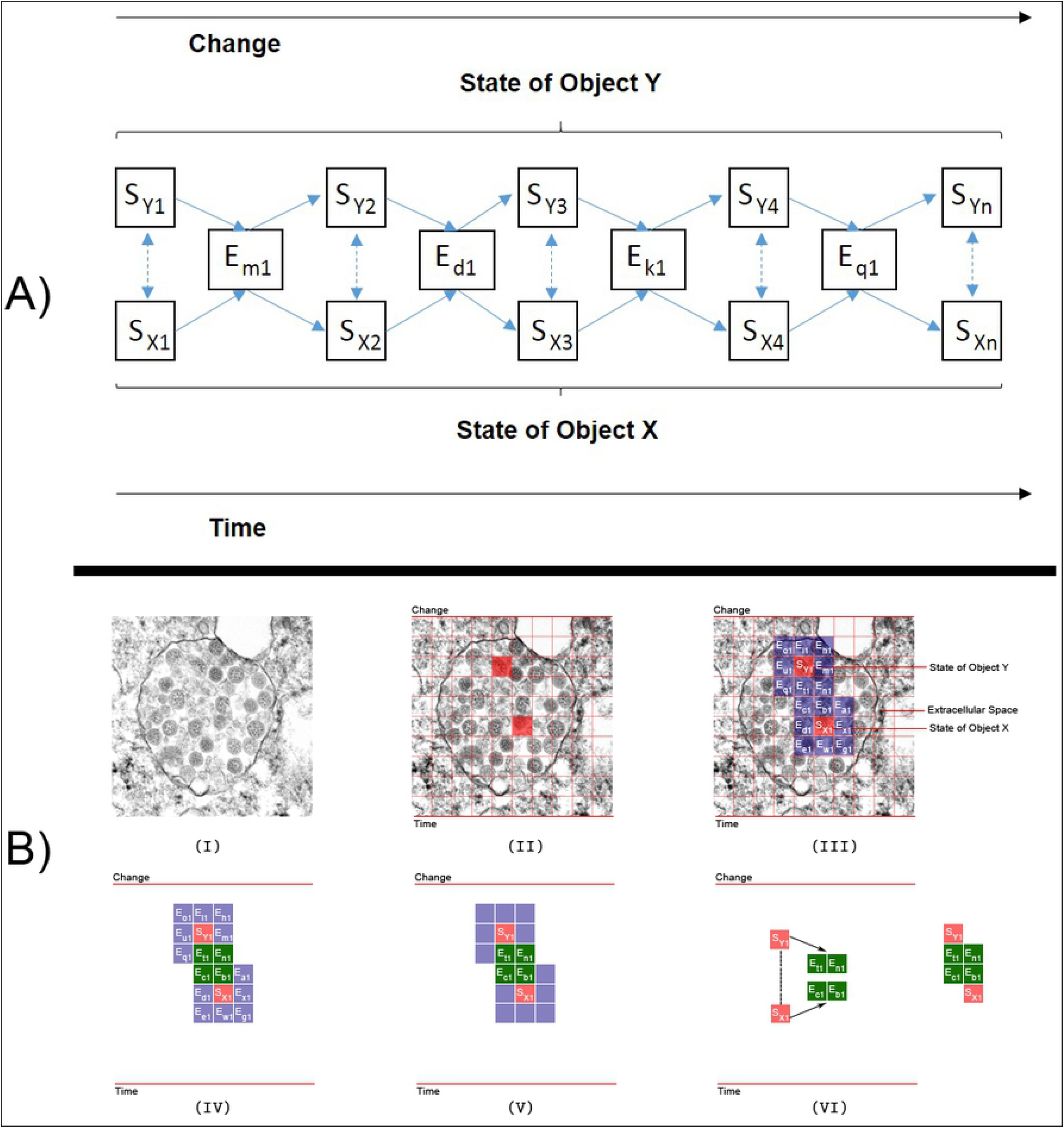
Extended framework version of COVNET45 for constructing and processing raster images.

The boxes labelled ”S” are states of those objects. Moreover, the boxes labelled ”E” are the events in which those objects participate, see figure 1.B.(III). The series numbers (1,2,3,n) in sets of boxes indicate sequences. The Y series in the top row of state boxes indicate states of object Y; similarly for the bottom row of object X and its states. The alphabetic on the event boxes indicate different types of events (Marked as Blue).

Each object can participate in different types of events (Marked as Red). The events of states Y and X that superseded with each other are the events of virus particles shedding (Marked as Green) see figures 1.B. (IV) and (V). The solid arrow shows each state of an object being superseded by a new state as the result of the object participating in the event. The double dashed-line arrows indicate states of objects that can affect and be affected by other states of objects see figure 1.B.(VI). Moreover, some objects can take part in events without being changed. For instance, an object in specific events can cause other objects to be changed.

The diagram of COVNET45 in figure (A) shows that only current states of objects can affect and be affected by other states. Past states of objects do not affect anything, except through their effect, directly or indirectly, on current states. Future states of objects do not affect anything because future states do not yet exist, and what does not exist cannot affect anything (although, of course, the present anticipation of future states by sentient beings may affect current states of those sentient beings, and through them, the current states of other objects).

Change is what results from a process. It is what goes on ”inside” an event. This process is a process of state transition. Before an object changes, it is in a given ephemeral state. It remains in that ephemeral state until it changes. After it changes, it is in a new ephemeral state, which may or may not also be a temporary state that it has been in before. These changes are what goes on in the events that happen to the object. This process of state/change/new state occurs over and over again, from when the object begins to exist to when it no longer exists.

The next section explains the technique employed by COVNET45 in processing nearly any giving objects in 2D images such as microdroplets or virus particles. Later we show how COVNET45 can provide accurate measurements in different scale systems.

### Image Utilization

COVNET45 can process different types of images specifically raster image files, since raster image files contain pixel values, and saved in an image file with JPG, JPEG, GIF, and PNG extension formats. COVNET45 also supports WEBP, JPS, JFIF, CUR, BMP, JPE and SVG image formats. In COVNET45 all uploaded images will remain unmodified and retain their original format; however, COVNET45 appends them internally to an existing DOM element for graphical representation reasons.

Fig 1 shows the extended framework version of COVNET45 for constructing and processing microscopic images into the tool. In the image construction phase, a referent is a type realized in any giving raster image at a time. Every referent (image) consists of pixels and each row of pixels consists of values. A referent is an object or an event. Therefore every pixel is an object or an event. And so the set represented by any pixel is either object of a specific type of events. In the mathematics phase, every bit of pixels is calculated in any giving image once uploaded to COVNET45. It calculates the length of the image in pixels and scales different types of bar values in pixels, and the resulting value is represented either as a value of object or an event or both.

Fig 2 shows a demonstration of an original microscopic image employed using the extended framework version of COVNET45 for constructing and processing raster images. The microscopic sample image demonstrates both states of object Y and X (Marked in red) with ten grid system blocks and the size of the image is 737-pixel in width and 737-pixel in height. The original image extension type is ”TIF”, and the original dimension pixel size is 1461-pixel width by 1461-pixel height. The referent is focusing only on the objects (two virus particles). We use ten grid system applied in the image to provide an understanding of COVNET45 frameworks. In image construction phase the highlighted grids (State of object Y and X) in red demonstrate the size of a single block in pixels, which also contains a single virus particle size, the dimensions of both states Y and X are 73-pixel widths by 73-pixel height.

**Fig 2.**
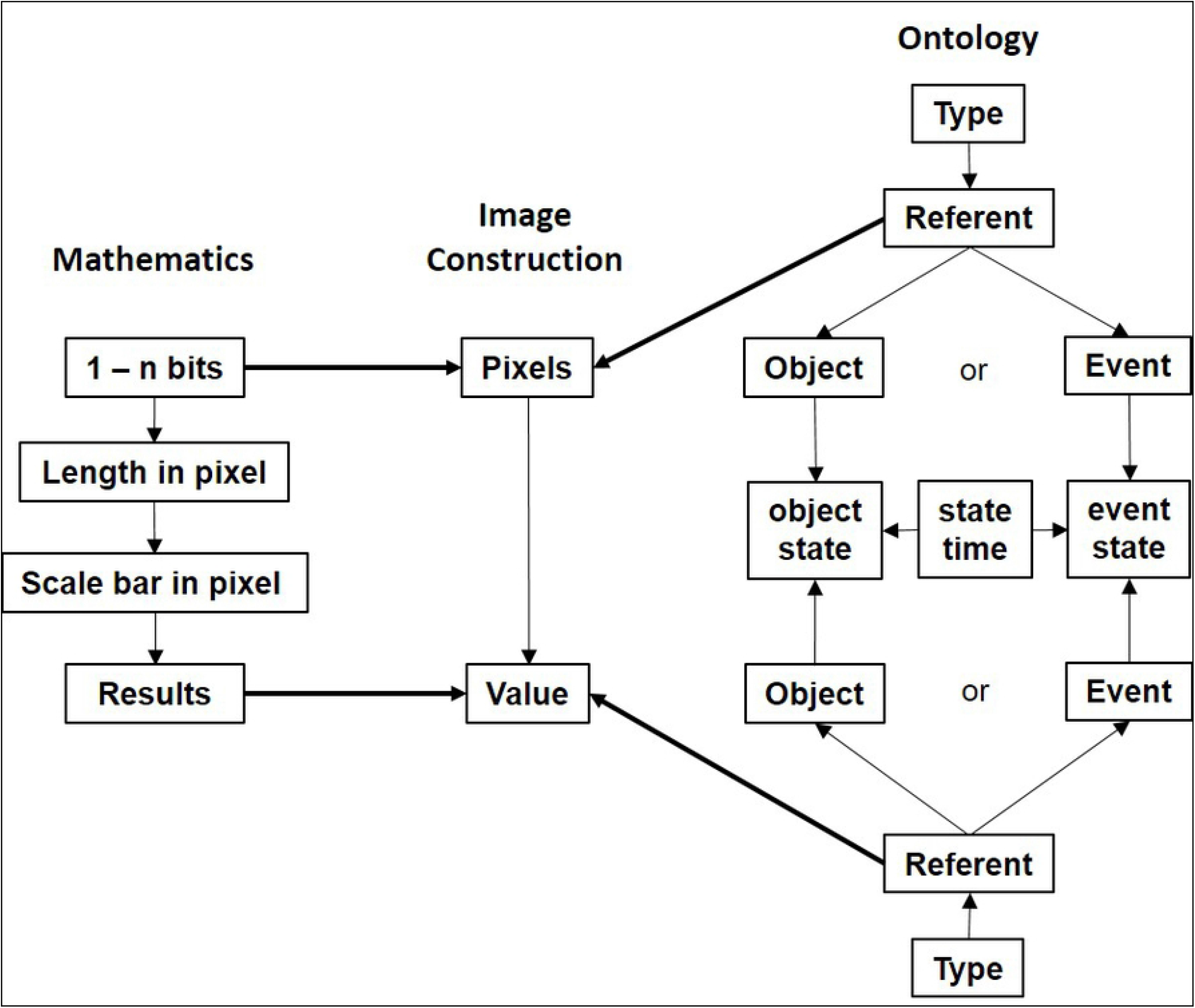
Sample microscopic image of COVID-19 virus, (a) Represent the pixel dimension of the image of 737×737 pixels highlighted with states of objects X and Y, (b) Shows the referent process of the extended framework of processing only object states of X and Y, (c) Shows the pixel value of objects X and Y explaining the image construction phase, (d) Shows the mathematics phase of measuring only the dimension of the virus (objects) in states X and Y.

In COVNET45, we calculate the size of many objects’ states; in this, we measure the size of a single virus particle in both states (Y and X). The size of the virus in object Y is 52.01-pixel in height. In contrast, the size of the virus in object X is the 37.01-pixel in height. Note that these measurements will be altered if an image has been resized or reshaped. In COVNET45, we use multilayer processing points in each uploaded image file. A user can construct and process raster images independently for a single image at a time. Fig 3 explains the technique used in COVNET45 for generating a new image, and the processing is described in details below.

**Fig 3.**
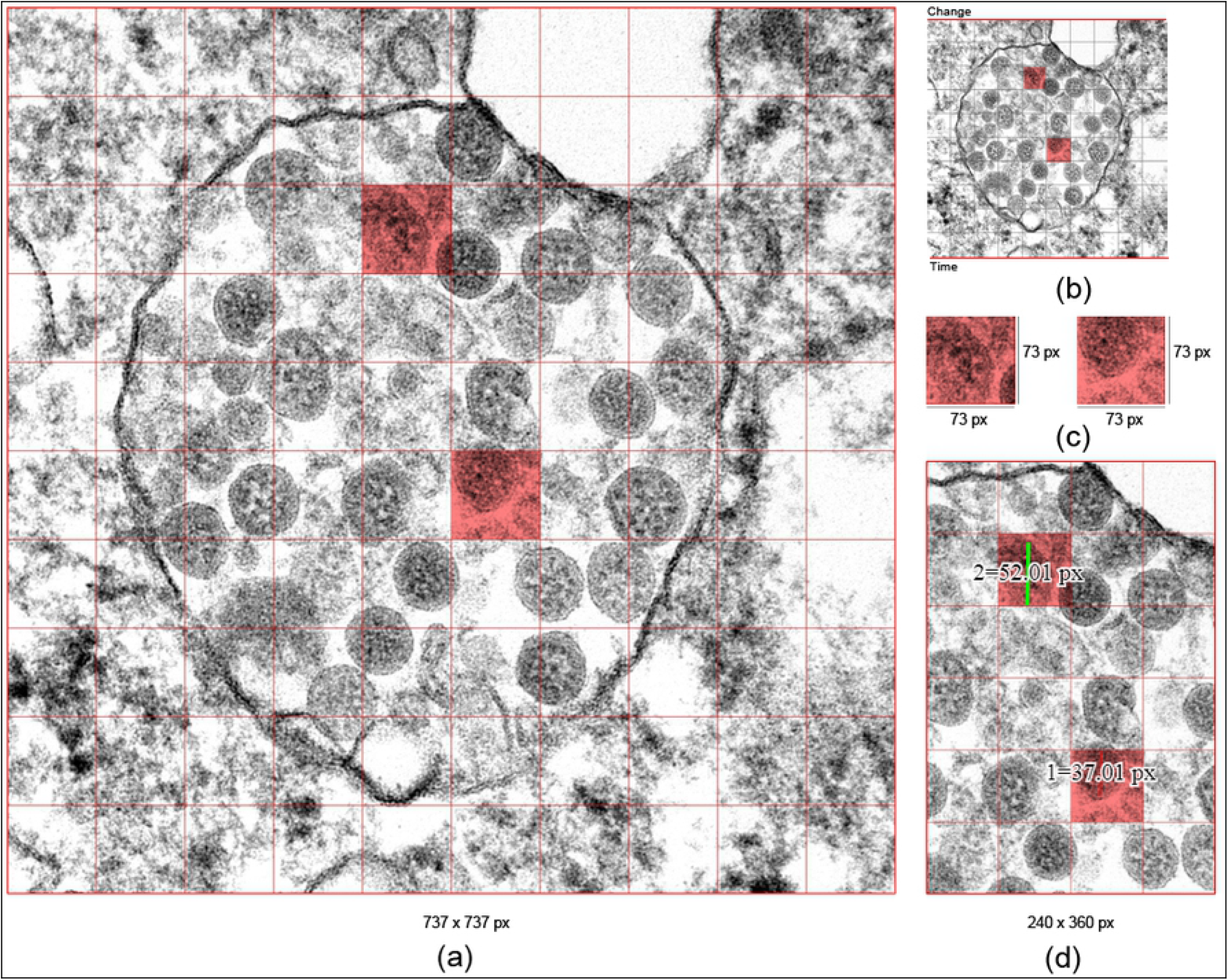
COVNET45 methods for processing raster images.

#### 1. Semantic Processing

The image preprocessing step is performed to process a single raster image uploaded to COVNET45 tool at a time. The left-hand side of the solution typically is the front end or channel referred to as (Image Type), this is strictly applied only to raster image format type of (JPG, JPEG, GIF, PNG, WEBP, JPS, JFIF, CUR, BMP, JPE, SVG). The brain behind COVNET45 would be the multi-layered automation functionality services. At the core of COVNET45, it provides automation solutions, which calculate measurements solution. Currently, COVNET45 supports five processing automation techniques described below.

- Image Segmentation (SEGBON): This automated feature uses an image segmentation technique adopting watershed algorithm, which provides an alteration on grayscale images to view it as a topographic surface. We present the watershed algorithm by using OpenCV programmed module adopted as a marker-based watershed transformation; every image pre-processed using this algorithm will be executed as a topographic surface.
- OpenCV Edge Detection (EDGE): This automated feature uses John F. Canny, canny edge detection algorithm, which is sensitive to noise that is eliminated using Gaussian filters to detect edges in the blurred image.
- Canny Edge Detection (CED): This automated feature uses a supervised version of OpenCV Edge Detection. Which is an improvement to the previous technique supervised CED introducing thresholds that can be added to a fine-tuned version of OpenCV Edge Detection, that are represented in sigma, thresholds, and Non-maximum suppression.
- Floyd-Steinberg Image Dithering (DITHER): This automated feature demonstrates an error-diffusion technique, intended to take advantage of binary images to increase the visual quality of the produced binary images. Where its calculations are based on quantization error between the pixel and its neighbouring pixels when scanned neighbours of scanned pixels are divided by error diffusion filter weights. The tool is taking the Floyd, Steinberg approach by using input image preprocessing as an over quantization then dithered as a grayscale output as a PNG compressed output, user interaction is prompt into selecting images without any selecting over algorithm weights.
- Image Optimization (OPTIZ): This automated feature similar to DITHER using Floyd, Steinberg algorithm. However, the only difference it allows altering the images in grayscale output, as well as allowing scaling the algorithm weights.

#### 2. Post Processing

The image post-processing step is performed after completion of the preprocessing step. This step consists of three stages, which are clarification, measurements and confirmation. Each image uploaded to COVNET45 will be processed using a user selection type of automation features described above. Once the output is confirmed, the users are eligible to perform the calculation of any selected area measurements within the image pixels.

To maintain a precise measurement scale of two points or more in each object in every state, whether, in state of object Y (Sy1) or state of object X (Sx1) during the post-processing stage, a right sampling approach over the states (X OR Y) had to be implemented. The main idea is to perform a universal conversion calculation mechanism to determine the precise sampling of each object within two or more points in a single image, and this will provide us with a significant verification of the size of objects in each image.

Definition 1 explains the measurements process. The definition represents the instances: G as green marker line, R as red marker line, M as calculated measurement per scale and S as the scale bar value. We assume that a single image consists of two markers to calculate a single object in a single uploaded image in COVNET45. Then COVNET45 generates a pixel value measurement for the red marker line and green marker line, which allows us to produce a measurements per scale. The resulting value of the measurements per scale when multiplied with the value of the integer scale bar value will result the accurate measurement of the object size within the same scale of the image used at time of upload to COVNET45.

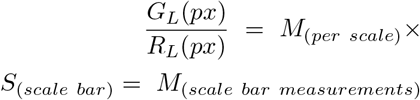

In Fig 6 (B) shows thin-section electron micrographs of the 2019 novel coronavirus grown in cells. We show the implementation of accurately measuring a single infected cell. Marked in green displacement diagonal line no. (2) and (3). After applying the formula, we can conclude the size of cell number (2) is 0.36 µm, while the size of the cell number (3) is 0.63 µm. Moreover, In Fig 4, we demonstrate and validate the virus’s particles estimated measurement, which is between 0.10 µm to 0.16 µm, which indicates that the virus particles size in an infected cell are approximated and can be measured correctly using electron micrographs microscope.

**Fig 4.**
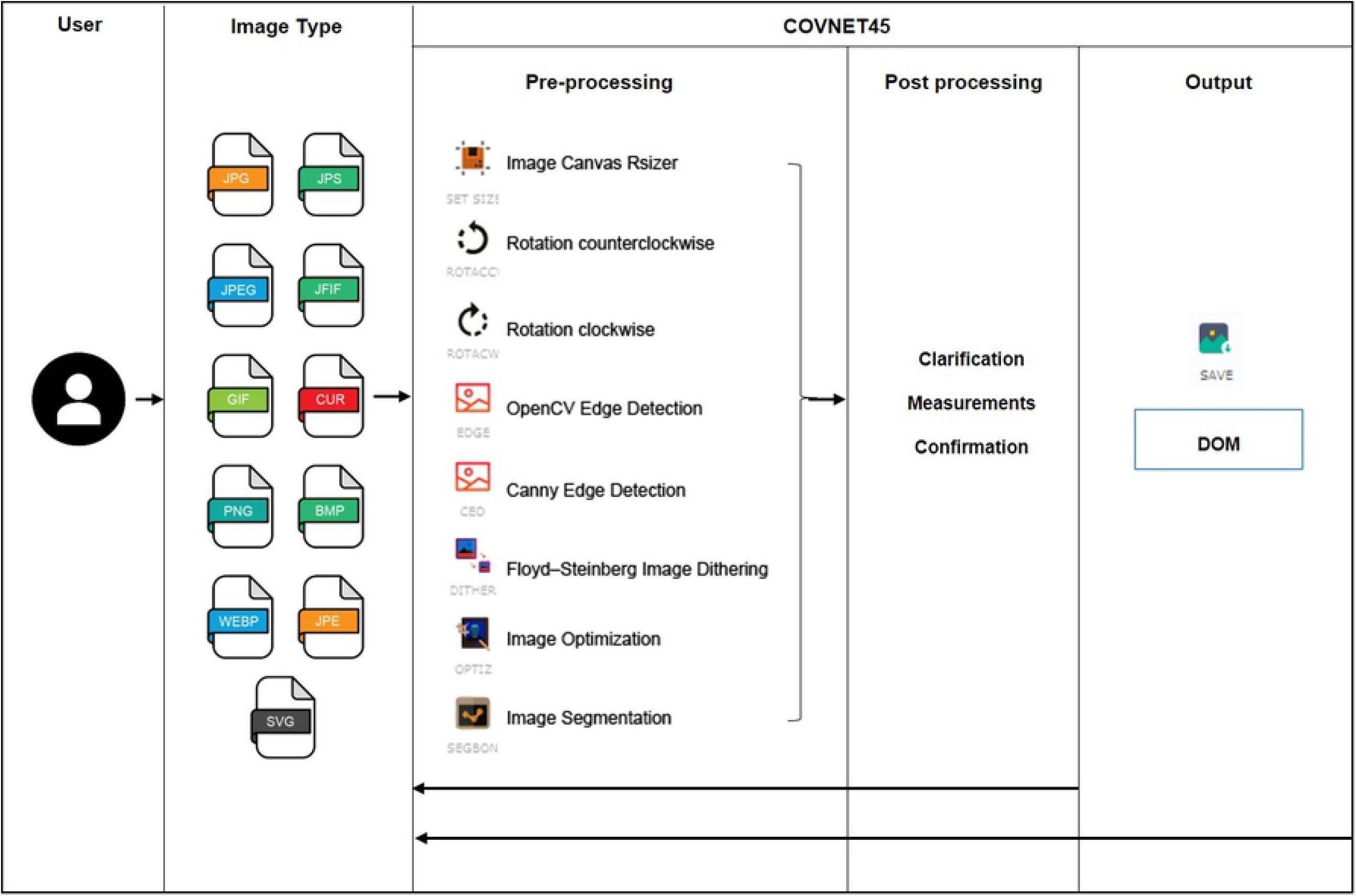
Original image shows thin-section electron micrograph of the 2019 novel coronavirus grown in cells published by the University of Hong Kong. The image shows part of a virus infected cell grown in culture with multiple virus particles being released from the cell surface.

In terms of measurements in different scale systems, the formula allows the scale to match the final results with the same image scale used in the photograph. In Fig 8 (C), we notice the image scale is on a centimetres scale. The research concludes that it corresponds to droplet size to nuclei of ca. 4 µm diameter, or 12-to 21-µm droplets before dehydration. Analyzing the highest droplet measurements in the image resulted in a different size than the research suggested. Marker sample number 2 considered the longest droplet in the image with a measurement size of 3.70 cm, and sample number 3 shows a tiny droplet with a measurement size of 0.23 cm. We conclude that COVNET45 uses the image scale, revealing the answer to the droplets’ correct measurements.

### Implementation and Experimental Evaluation

COVNET45 uses standard web technologies and is hosted on AWS cloud servers. The software presented as a cloud-based software architecture, Software-as-a-Service (SaaS), built with the latest Codeignitor V3 framework. Below we discuss the features of the experiments provided by COVNET45.

#### 1. Image Segmentation (SEGBON)

Fig 5 Demonstrates the results of two test images authenticating the use of Image Segmentation (SEGBON) approach in COVNET45. It shows the use of image segmentation approach over watershed transformation, which allows adding foreground extraction using the ”GrabCut” algorithm. It allows region outside the box boundaries of foreground first then segment the content within the box region, user interaction selecting objected to assigning them as a foreground or background [21].

**Fig 5.**
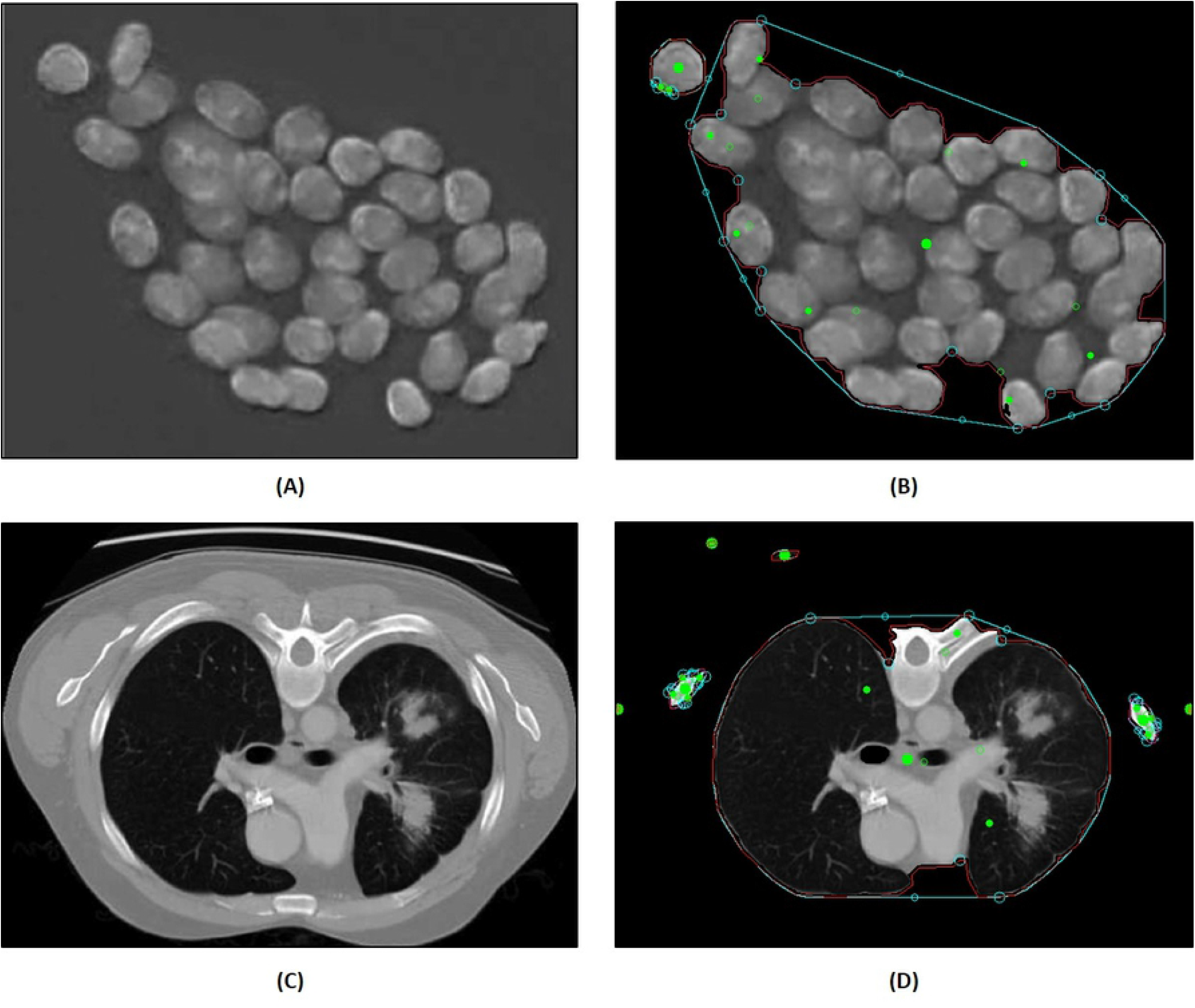
Demonstrates image segmentation (SEGBON)feature in COVNET45: A) The original image of hematoxylin-stained objects (nuclei) [23]. B)Shows the output image after applying segbon segmentation in COVNET45. C) Demonstrates The original input image of the vertical lung scan of automatic lung tumor segmentation on PET/CT images using fuzzy markov random field model [22]. D) Shows the output image after applying segbon segmentation in COVNET45.

In Figure (A) it shows the result in a foreground segmentation based on ”GrabCut” nuclei images by eliminating the box region. Using the original image in COVNET45 figure (B) the result shows that overlapped particles and spaces are not segmented by applying the mask, and boundaries have been judged based on the segmented object’s edges [23].

Figures (C) and (D) test image both results in revealing similarity in regions of interest since those regions are segmented similarly. This case can be generalized to similar cases in extracting segments of interest in similar medical images, where its iterative approach is segmenting the object by border matting around the rigid segmentation boundary [22].

#### 2. OpenCV Edge Detection (EDGE)

Fig 6 Demonstrates the results of single test image authenticating the use of OpenCV Edge Detection (EDGE) algorithm approach in COVNET45. Figure (A) shows the original image of thin-section electron micrograph of the 2019 novel coronavirus grown in cells, the image published by the University of Hong Kong. The image demonstrates part of the virus-infected cell grown in a culture with multiple virus particles being released from the cell surface.

**Fig 6.**
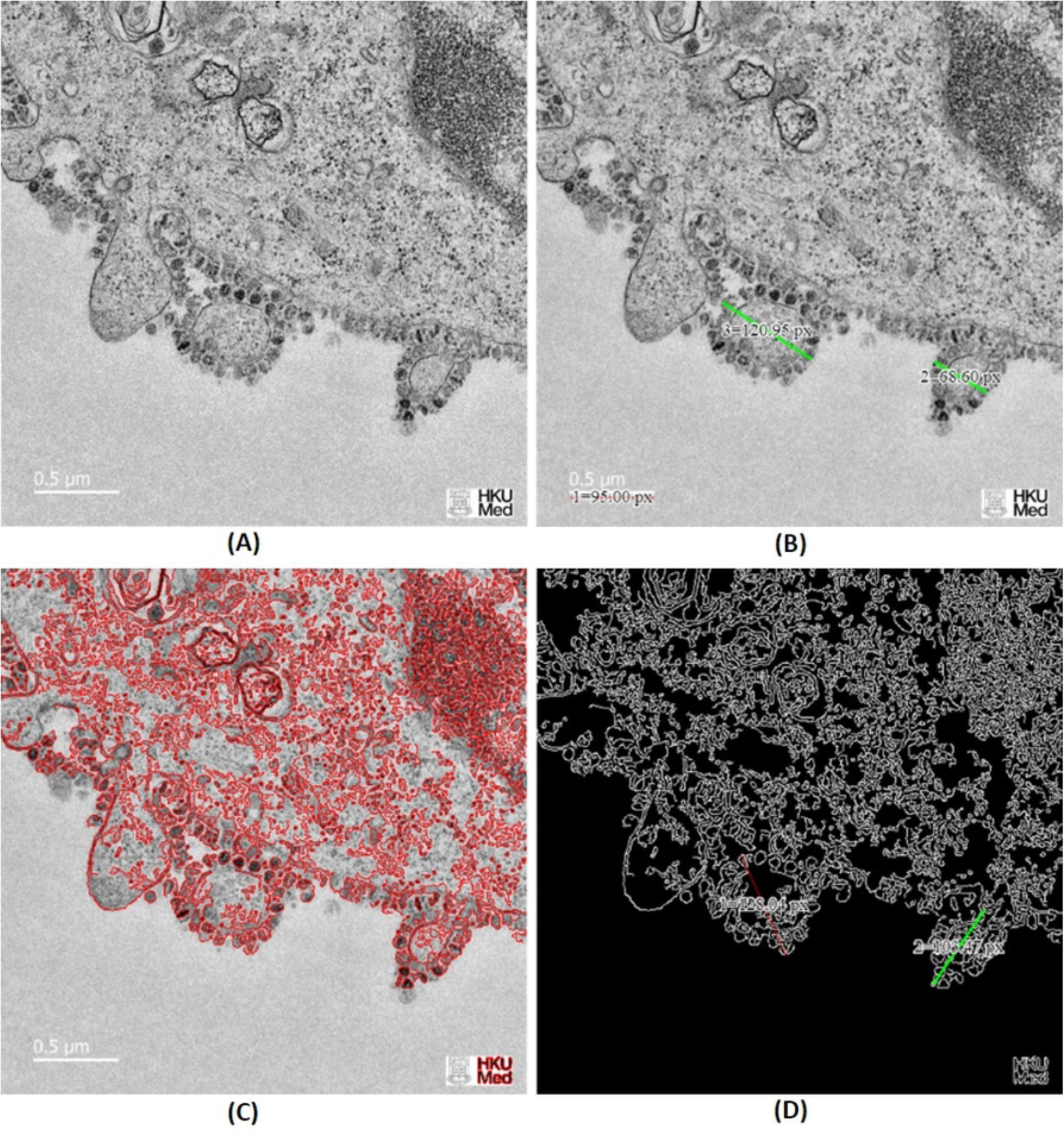
Demonstrates OpenCV Edge Detection (EDGE) feature in COVNET45: A) Original image shows thin-section electron micrographs of the 2019 novel coronavirus grown in cells published by the University of Hong Kong. The image shows part of a virus infected cell grown in culture with multiple virus particles being released from the cell surface. Each infected cell produces thousands of new infectious virus particles, which can go on to infect new cells. Image credit: John Nicholls, Leo Poon and Malik Peiris, LKS Faculty of Medicine, and Electron Microscopy Unit, The University of Hong Kong. B) Shows exported image marked with sample measurements in pixel to two infected cells with COVID-19 using COVNET45 tool. C)Shows the implementation of ridge detector to the original image in COVNET45 using the feature OpenCV Edge Detection (EDGE). D)Shows the implementation of canny edge detector to the original image in COVNET45 with the use of non-maximum suppression technique using the feature OpenCV Edge Detection (EDGE).

Moreover, figures (C) and (D) demonstrate the technique used in COVNET45 using ridge detector and canny edge detector, revealing more information in the image edges than the original image. Detecting edges can help measure cell with higher precision compared to the original image.

#### 3. Canny Edge Detection (CED)

Similarly, Fig 7 demonstrate the feature Canny Edge Detection (CED) which uses the same technique as the OpenCV Edge Detection (EDGE) technique. However, the regions in any uploaded image are more automated in Canny Edge Detection (CED) depending on the threshold provided when uploading the image. Canny Edge Detection (CED) can refine extra details to the images by the user based thresholds, which help reveal the darker shades of dark regions in the image. The marked regions 2 and 3 are examples of applied thresholds to reveal the cells’ details, so the measure tool can be applied to measure the length of interest cells.

**Fig 7.**
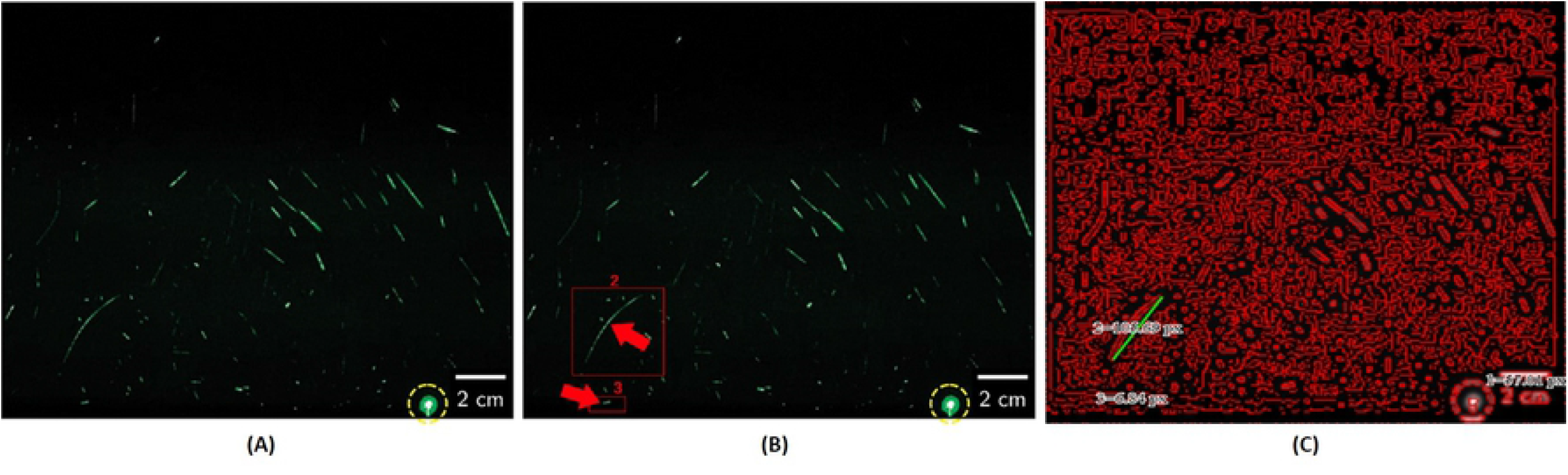
Demonstrates Canny Edge Detection (CED) feature in COVNET45: A) Original image shows speech droplets using highly sensitive laser light, the image describe the observation of airborne speech droplet nuclei, generated by a 25-s burst of repeatedly speaking the phrase “stay healthy” in a loud voice, the droplet size to nuclei is 4 µm diameter, or 12-to 21-µm droplets prior to dehydration. B) The marker demonstrates the test measurements that COVNET45 will verify. C) Shows iteration of the image using the canny edge detection (CED) technique in COVNET45, which reveals the edges of the droplets using high intensity thresholds. The real measurements of the sample number 2 considered the longest droplet in the image with the measurement size of 3.70 cm and sample number 3 shows a small droplet with measurement size of 0.23 cm.

#### 4. Floyd-Steinberg Image Dithering (DITHER)

Fig 8 demonstrates the feature Floyd-Steinberg Image Dithering (DITHER). Figure (A) shows the original image recording of a real person coughing as a test subject. It demonstrates that air droplets can travel 1 m away from the mouth [19]. Figure (B) demonstrates the original image using the Floyd Steinberg algorithm distortion to generate additional details on the image. This technique used in COVNET45 to add extra sharpness to the image, which reveals more details of the air droplet particles. Added sharpness in dithering reduces the error artefacts when low-quality images are input to the tool, allowing the recovery of the information lost to under-sampling by pixel in images that can be used to recover the details of nearly any imported images into COVNET45.

**Fig 8.**
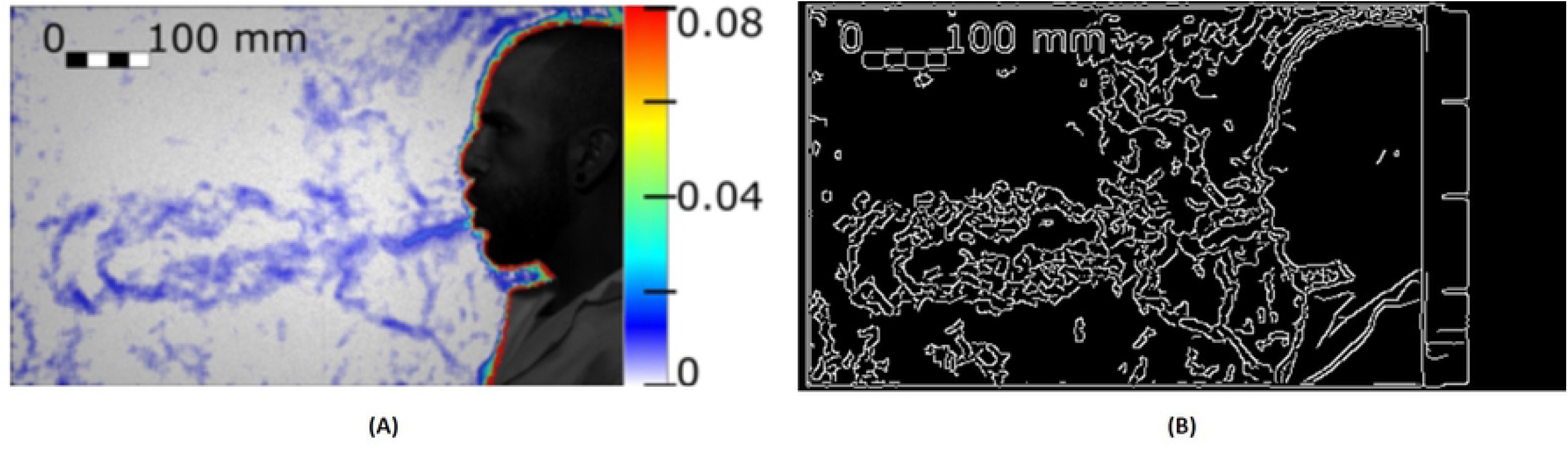
Demonstrates Floyd-Steinberg Image Dithering (DITHER) feature in COVNET45: A) Original image shows the test of a real person coughing, which generates an airflow. The image was taken using a high-speed CMOS camera (VEO710L, Vision Research) with 1280 × 800 square pixel was used for recording the displacement of the droplets is measured as 1 meter away from the mouth [19]. B) Shows exported image using Error diffusion using the Floyd Steinberg dithering algorithm using COVNET45 tool, revealing more characteristics of air droplets particles in the image.

#### 5. Image Optimization (OPTIZ)

Fig 9 demonstrate the feature Image Optimization (OPTIZ). Figure (A) shows representative fluorescent micrograph of fibrous or cellular deposits in the plasma smears of COVID-19 patients [20]. This feature result is similar to error diffusion using Floyd-Steinberg Image Dithering (DITHER) algorithm. This feature allows the users to customize the image from background colours and delete certain features from the uploaded image. Uploaded images into this feature provide transparency to the background, which highlights the objects in the images. Exported images using OPTIZ can be stored at either black and white, inverted black and white in 2, 4 or 8 bits per pixel (bpp).

**Fig 9.**
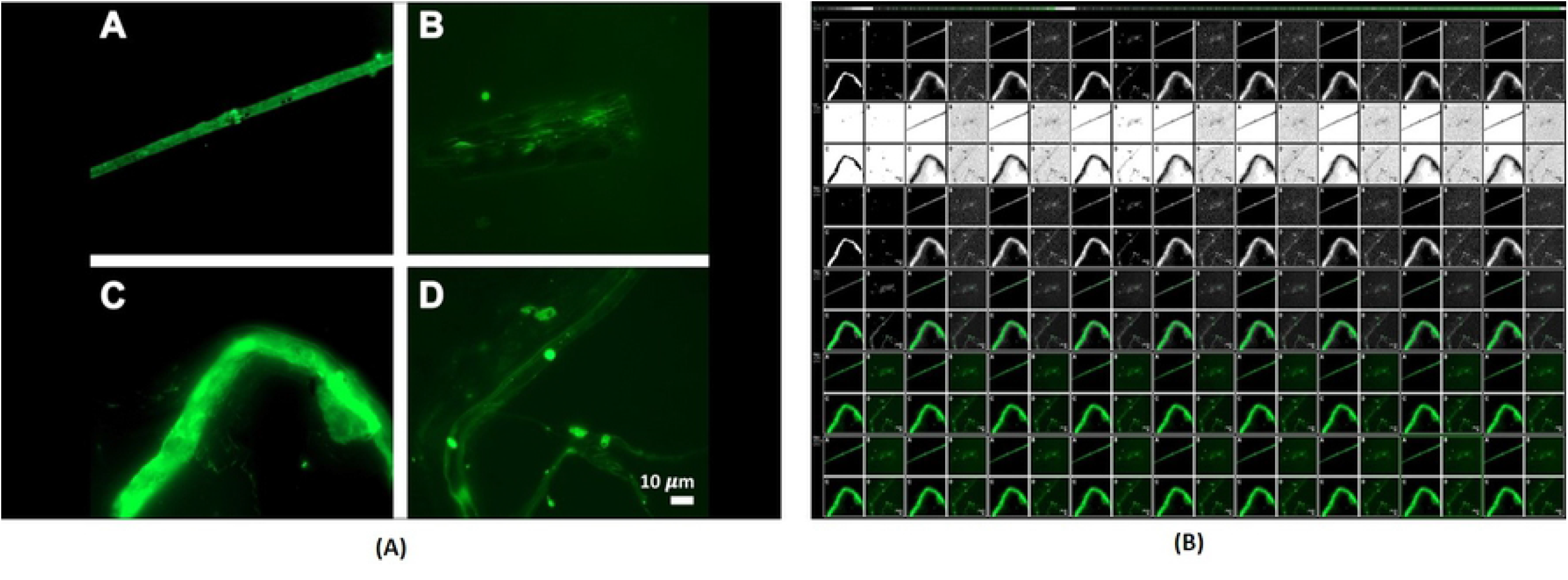
Demonstrates Image Optimization (OPTIZ) feature in COVNET45: A) Original image shows representative fluorescent micrographs of fibrous or cellular deposits in the plasma smears of COVID-19 patients. The image was taken from the research prevalence of amyloid blood clots in COVID-19 plasma [20]. B) Shows the exported images using Image Optimization (OPTIZ) in COVNET45 tool.

In order to test the tool, we used several images published in different research as seen in Fig 2, Fig 4, Fig 5, Fig 6, Fig 7, Fig 8 and Fig 9. The experimentation and development are carried on Fujitsu Siemens Lifebook laptop model AH532/G21, with 8GB of memory, 2.6 GHz Intel Core i5 processor with HD graphics 4000 Nvidia Geforce. The web application is built as SaaS hosted in AWS cloud with AES 256 encryption for added privacy. COVNET45 tool was tested using Google Chrome version 88 and Firefox version 72. COVNET45 is coded using the Application Development Framework (CodeIgnitor V.4) using PHP and MySQL 5.1+ as a database structure.

COVNET45 synthesis a robust solution to optimize and measure a variety of medical and non-medical raster images types. In Fig 10 we show COVNET45 dashboard and application screens in responsive view and web view. COVNET45 was able to authenticate and measure high accuracy measurement results by optimizing raster image types. COVNET45 combines strong capabilities of optimizing raster images with automation detection mechanisms and accurately measuring different scale systems. Furthermore, COVNET45 handles only a single image as an input terminal upload and limited to the raster image type of JPG, JPEG, GIF, and PNG extension format. COVNET45 also supports WEBP, JPS, JFIF, CUR, BMP, JPE and SVG image formats. Nonetheless, the tool has the following limitations.

**Fig 10.**
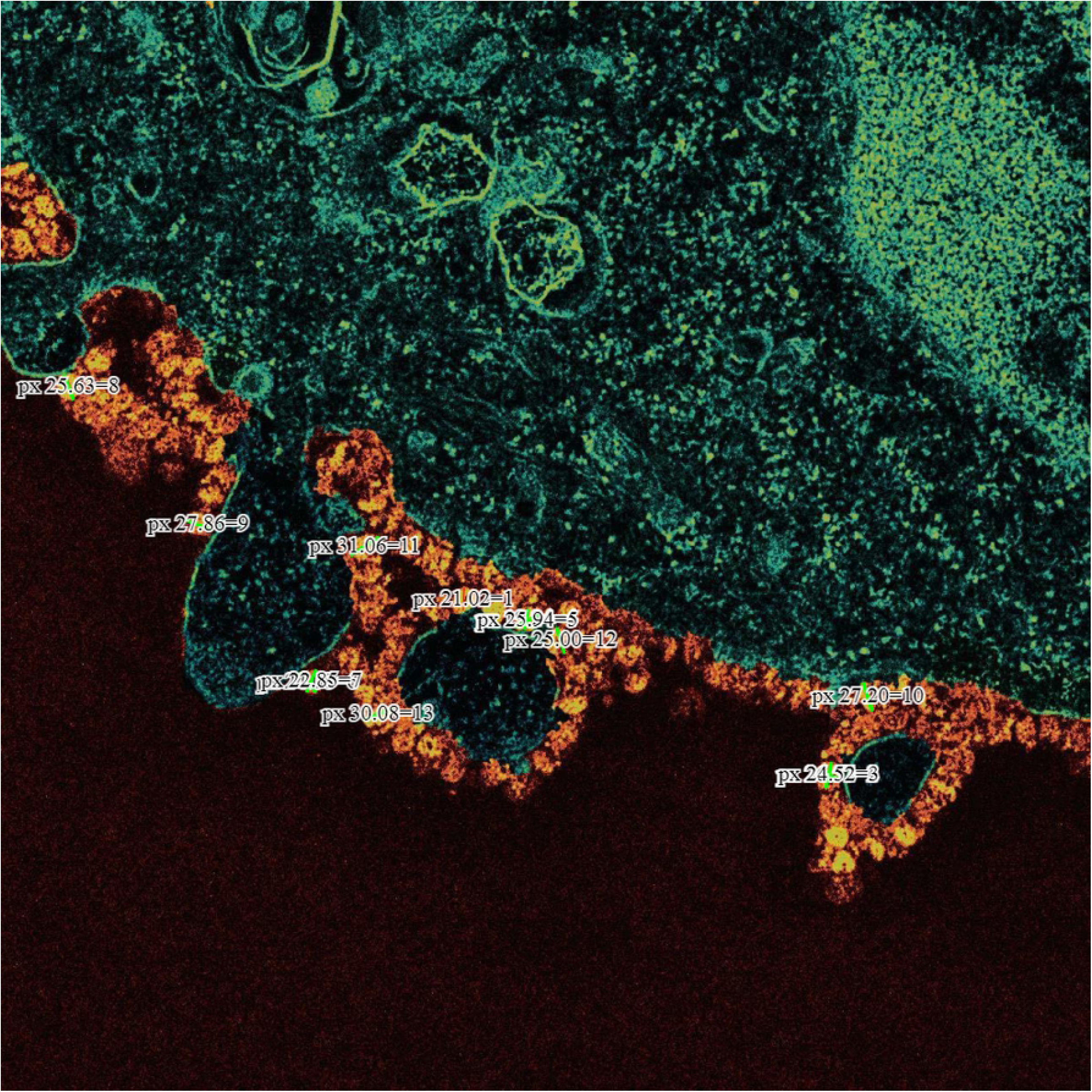
COVNET45 dashboard and application screens in responsive and web views. The screens demonstrate the machine learning algorithms screens and the control panel for administration access privileges.

- Image segmentation feature (SEGBON) unable to process certain objects if the image’s foreground is not completely black, therefore the algorithm won’t distinguish small particles that contain a similar colour of the image foreground.
- COVNET45 uses different machine learning algorithms in each feature, which may slow automation, that is done in the cloud and processed in the client DOM browser.
- COVNET45 uses images scale bars to measure accurately any giving object in the image; without the scale system applied in the image, COVNET45 won’t be able to distinguish the level scale system to be used.

Moreover, respiratory transmission depends on the incorporation of the airborne particles in aerosols. Aerosols are produced during speaking and regular breathing, while coughing produces even more forceful expulsion. Transmission from the nasal cavity is facilitated by sneezing and is much more effective if the infection induces nasal secretions. A sneeze produces up to 20,000 droplets (in contrast to several hundred expelled by coughing), and all may contain rhinovirus if the individual has a common cold. As noted when we discussed viral entry, the size of a droplet affects its ”hang time”: large droplets fall to the ground, but smaller droplets (1 to 4 um in diameter) may remain suspended in the air for a longer time. Nasal secretions also frequently contaminate hands or tissues. The infection may be transmitted when these objects contact another person’s fingers, and that person, in turn, touches his or her nose or conjunctiva. In today’s crowded cities, transport and workplaces, people’s physical proximity may facilitate viruses to spread more effectively. In Fig **??** we show how COVNET45 would be able to detect events of particles spread in cells, where a single spherical virus particles contain black dots of COVID-19 virus as an *object* in a single microscopic image, and the possible movements/diameter in size of a single spherical virus particle as *events*. Spherical virus particles contain black dots, which are cross-sections through the viral nucleocapsid. In the cytoplasm of the infected cell, clusters of particles are found within the membrane-bound cisternae of the rough endoplasmic reticulum/Golgi area. This will allow us to develop a pattern recognition model solution and behaviour movement of spread in micrograph images and cells infection that can be measured before virus spread in the human body.

## CONCLUSION AND FUTURE WORK

This paper presented an automatic measurement detection tool, ”COVNET45”. The tool provides several automatic intelligent techniques to process, detect and measure any giving objects and events in raster image files highlighted in this paper with a discussion of microdroplet transmissions in airborne space. COVNET45 will allow scientists to automatically optimize raster images, such as micrograph images of microscopic images, and perform precise measurements observation to any given object or particles.

The experimental results also demonstrated that the COVNET45 is helpful in processing high-resolution images and exploring detailed or hidden characteristics, allowing users to have complete control of any uploaded image of their choice. Biologist uses microscopic to study biological data to analyse cellular structure, organism and characteristics. Understanding different cell characteristic under different conditions like germ, viruses and effect of cell proliferation to examine the other conditions in-depth using AI algorithmic software’s. COVNET45 can examine anatomical cell and measure characteristics like capturing, tracking cells like microscopic studies a common tool as ImageJ software that is an addition to use in parallel. However, work is continuous to allow users to have multiple uploads rather than a single upload to the system and add more graphical features to control images. Providing markup comments and highlighted select into the images. Finally, we are working on developing an internal mechanism to minimize external calls to third-party modules. COVNET45 is being integrated with DOM scripting, eliminating uploads and not storing images in the server, which provide efficiency in image optimization automation. COVNET45 is currently hosted in AWS and can be integrated with the AMAZON AI machine learning platform. This will allow the tool to transmit and perform optimization to the images more effectively and enhance security.

## Data Availability

COVNET45 is currently hosted in AWS and can be integrated with the AMAZON AI machine learning platform Tool, Data and code will be shared via https://github.com/HeshamAlsaadi

## Acknowledgment

This work was supported by Zayed University Research Office, Research Incentive Fund Grant #R20089

---

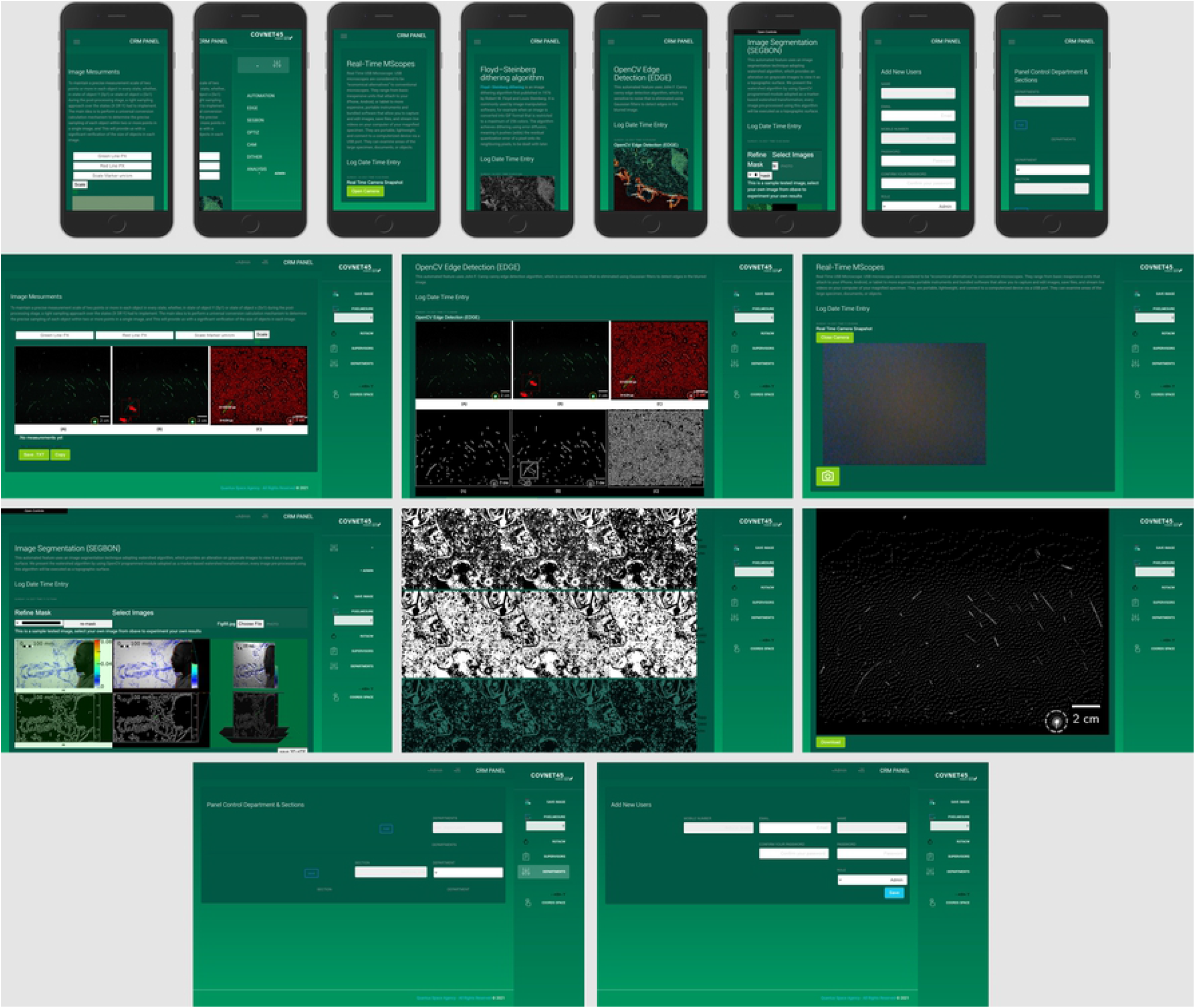

## References

1. Wu, D., Lu, J., Liu, Y., Zhang, Z., & Luo, L. (2020). Positive effects of COVID-19 control measures on influenza prevention. International Journal of Infectious Diseases, 95, 345–346.

2. Lee, J., Yoo, D., Ryu, S., Ham, S., Lee, K., Yeo, M., … & Yoon, C. (2019). Quantity, size distribution, and characteristics of cough-generated aerosol produced by patients with an upper respiratory tract infection. Aerosol and Air Quality Research, 19(4), 840–853.

3. Stadnytskyi, V., Bax, C. E., Bax, A., & Anfinrud, P. (2020). The airborne lifetime of small speech droplets and their potential importance in SARS-CoV-2 transmission. Proceedings of the National Academy of Sciences, 117(22), 11875–11877.

4. Asadi, S., Bouvier, N., Wexler, A. S., & Ristenpart, W. D. (2020). The coronavirus pandemic and aerosols: Does COVID-19 transmit via expiratory particles?. Aerosol Science and Technology, 54(6), 635–638.

5. Duguid, J. P. (1946). The size and the duration of air-carriage of respiratory droplets and droplet-nuclei. Epidemiology & Infection, 44(6), 471–479.

6. Morawska, L. J. G. R., Johnson, G. R., Ristovski, Z. D., Hargreaves, M., Mengersen, K., Corbett, S., … & Katoshevski, D. (2009). Size distribution and sites of origin of droplets expelled from the human respiratory tract during expiratory activities. Journal of aerosol science, 40(3), 256–269.

7. Gralton, J., Tovey, E., McLaws, M. L., & Rawlinson, W. D. (2011). The role of particle size in aerosolised pathogen transmission: a review. Journal of Infection, 62(1), 1–13.

8. Van Doremalen, N., Bushmaker, T., Morris, D. H., Holbrook, M. G., Gamble, A., Williamson, B. N., … & Munster, V. J. (2020). Aerosol and surface stability of SARS-CoV-2 as compared with SARS-CoV-1. New England journal of medicine, 382(16), 1564–1567.

9. Machida, M., Nakamura, I., Saito, R., Nakaya, T., Hanibuchi, T., Takamiya, T., … & Inoue, S. (2020). Adoption of personal protective measures by ordinary citizens during the COVID-19 outbreak in Japan. International Journal of Infectious Diseases, 94, 139–144.

10. Douglas, D., & Douglas, R. (2020). Addressing the corona virus pandemic: will a novel filtered eye mask help?. International Journal of Infectious Diseases, 95, 340–344.

11. Yunus, A. P., Masago, Y., & Hijioka, Y. (2020). COVID-19 and surface water quality: Improved lake water quality during the lockdown. Science of the Total Environment, 731, 139012.

12. Selvaraj, G., Kaliamurthi, S., Peslherbe, G. H., & Wei, D. Q. (2021). Are the Allergic Reactions of COVID-19 Vaccines Caused by mRNA Constructs or Nanocarriers? Immunological Insights. Interdisciplinary Sciences: Computational Life Sciences, 1–4.

13. World Health Organization. (2020). Overview of public health and social measures in the context of COVID-19: interim guidance, 18 May 2020 (No. WHO/2019-nCoV/PHSM Overview/2020.1). World Health Organization.

14. Ningthoujam, R. (2020). COVID 19 can spread through breathing, talking, study estimates. Current medicine research and practice, 10(3), 132.

15. Karim, M., Döhmen, T., Rebholz-Schuhmann, D., Decker, S., Cochez, M., & Beyan, O. (2020). Deepcovidexplainer: Explainable covid-19 predictions based on chest x-ray images. arXiv preprint 2004.04582.

16. Ai, T., Yang, Z., Hou, H., Zhan, C., Chen, C., Lv, W., … & Xia, L. (2020). Correlation of chest CT and RT-PCR testing for coronavirus disease 2019 (COVID-19) in China: a report of 1014 cases. Radiology, 296(2), E32–E40.

17. Verma, S., Dhanak, M., & Frankenfield, J. (2020). Visualizing the effectiveness of face masks in obstructing respiratory jets. Physics of Fluids, 32(6), 061708.

18. Blocken, B., Malizia, F., Van Druenen, T., & Marchal, T. (2020). Towards aerodynamically equivalent COVID19 1.5 m social distancing for walking and running. Preprint.

19. Viola, I. M., Peterson, B., Pisetta, G., Pavar, G., Akhtar, H., Menoloascina, F., … & Mehendale, F. V. (2021). Face coverings, aerosol dispersion and mitigation of virus transmission risk. IEEE Open Journal of Engineering in Medicine and Biology, 2, 26–35.

20. Pretorius, E., Venter, C., Laubscher, G. J., Lourens, P. J., Steenkamp, J., & Kell, D. B. (2020). Prevalence of amyloid blood clots in COVID-19 plasma. medrxiv.

21. Rother, C., Kolmogorov, V., & Blake, A. (2004). ” GrabCut” interactive foreground extraction using iterated graph cuts. ACM transactions on graphics (TOG), 23(3), 309–314.

22. Sun, S., Bauer, C., & Beichel, R. (2011). Automated 3-D segmentation of lungs with lung cancer in CT data using a novel robust active shape model approach. IEEE transactions on medical imaging, 31(2), 449–460.

23. Kowal, M., Żejmo, M., Skobel, M., Korbicz, J., & Monczak, R. (2020). Cell nuclei segmentation in cytological images using convolutional neural network and seeded watershed algorithm. Journal of digital imaging, 33(1), 231–242.

24. Smereka, J., Ruetzler, K., Szarpak, L., Filipiak, K. J., & Jaguszewski, M. (2020). Role of mask/respirator protection against SARS-CoV-2. Anesthesia and analgesia.

